# Assessment of the potential use of VAL-1221 for Lafora disease: MS-based proteomics for the characterization and quantitation of the biotechnological drug in plasma and cerebrospinal fluid

**DOI:** 10.1101/2025.09.17.25335891

**Authors:** Erika Esposito, Alice Caravelli, Lorenzo Muccioli, Chiara Cancellerini, Maria Tappatà, Eleonora Pizzi, Raffaella Minardi, DEFEAT-LD Study Group, Valerio Carelli, Luca Vignatelli, Roberto Michelucci, Francesca Bisulli, Jessica Fiori

**Author notes:** **Corresponding author:** Lorenzo Muccioli, MD, PhD, IRCCS Istituto delle Scienze Neurologiche di Bologna, Via Altura 1/8, 40139 Bologna - Italy. DEFEAT-LD study group: a list of collaborators is provided at the end of the manuscript as APPENDIX.

## Abstract

**Background:** VAL-1221 is a biotechnological fusion protein that combines the Fab portion of a cell-penetrating antibody with recombinant human acid α-glucosidase. Originally used for the treatment of Pompe disease, it has since attracted interest for possible repurposing in Lafora disease (LD), an ultra-rare, fatal form of progressive myoclonus epilepsy characterized by the accumulation of polyglucosan aggregates (Lafora bodies, LBs) within the central nervous system (CNS). Given its design, which includes a cell-penetrating domain, VAL-1221 has been hypothesized to cross the blood–brain barrier (BBB) and target pathogenic glycogen deposits within the CNS in LD. This study aimed to investigate the presence of VAL-1221 in plasma and cerebrospinal fluid (CSF) of LD patients and assess its potential to cross the BBB, using high-resolution mass spectrometry coupled with micro-liquid chromatography (microLC-HRMS/MS).

**Methods:** As part of a compassionate use program, five LD patients received intravenous VAL-1221 (20 mg/kg, every other week). LD untreated patients were included as controls. Plasma samples were collected at multiple time points up to 24 hours post-infusion, and CSF samples were obtained based on concentration profiles. Untargeted-to-targeted bottom-up proteomics were used to detect a unique peptide tag in biological fluids. Method validation included assessments of precision, accuracy, matrix effects, and analyte stability.

**Results:** VAL-1221 was consistently detected in plasma up to 4 hours post-infusion, while no VAL-1221 was detected in CSF samples with the method’s limit of detection. The validated method showed high sensitivity, precision (RSD ≤15%), accuracy (RE ≤15%), and acceptable matrix effect. Recovery was optimal for CSF; it was low in plasma (Rec% > ±20).

**Conclusion:** VAL-1221 was reliably detected in plasma after infusion, but no measurable levels were observed in CSF based on the validated method’s sensitivity. These findings suggest that the drug, when administered intravenously, may not reach the central nervous system, indicating that this route may not be appropriate for efficacy.

## 1. Introduction

Biopharmaceutical drugs represent a unique class of medicines due to their complex molecular structure. The growing biopharmaceutical market is driven mainly by monoclonal antibodies (mAbs), bispecific mAbs, antibody-drug conjugates (ADCs), antigen-binding fragments of mAbs (Fab), and proteins linked to the fragment crystallizable (Fc) region of mAbs (Fc-fusion proteins) [1]. In contrast to conventional small-molecule drugs, which typically have masses not exceeding a few hundred Daltons, biotechnological therapeutics are significantly larger, ranging from 5 kDa (e.g., insulin) to well over 100 kDa. These drugs exhibit structural complexity that can significantly influence their biological activity, therapeutic efficacy and toxicity, necessitating comprehensive characterization.

VAL-1221 is an investigational biotechnological drug produced by Parasail LLC as a treatment for patients with Pompe disease, a glycogen storage disease type II. VAL-1221’s glycogen-targeting approach has also prompted interest in conditions like Lafora disease (LD), where abnormal glycogen plays a key role. LD is an ultra-rare, genetically inherited (autosomal recessive), fatal form of progressive myoclonus epilepsy with adolescent onset. It is characterized by the accumulation of poorly branched, insoluble glycogen-like polymers, known as Lafora bodies (LBs), primarily within neurons and other cells of the central nervous system (CNS) [2,3]. Currently, there are no approved treatments beyond symptomatic management and supportive care.

From a structural standpoint, VAL‐1221 is a fusion protein of 147 kDa, combining the Fab fragment of a cell-penetrating antibody (3E10) with recombinant human acid α‐glucosidase (rhGAA) [4].

While conventional enzyme replacement therapies such as Myozyme^®^ and Lumizyme^®^ rely on mannose-6-phosphate receptor (M6PR)-mediated uptake to deliver GAA to lysosomes [5], VAL-1221 is engineered to access both lysosomal and cytoplasmic glycogen. This is particularly relevant in Lafora disease, where the pathological aggregates (LBs) accumulate predominantly in the cytoplasm of neurons [3]. The Fab domain of 3E10 facilitates cellular entry through the ENT-2 nucleoside transporter, a mechanism that has also been associated with translocation across brain endothelial cells in in vitro models of the blood–brain barrier [6]. Recent research using LD animal models has demonstrated that delivering a polyglucosan-degrading antibody-enzyme fusion directly to the brain can clear LBs and improve neurological manifestations [7]. Still, the current VAL-1221 formulation is not suitable for human ICV injections. Thus, if intravenously administered, VAL-1221 can cross the BBB, it might be able to break down cerebral LBs and represent a promising therapeutic strategy for patients with LD.

The development of biotechnological drugs for CNS disorders is particularly challenging due to the restrictive nature of the BBB, which limits the passage of macromolecules, including therapeutic proteins. Various strategies have been explored to enhance CNS drug delivery, including receptor-mediated transcytosis [8], nanoparticle-based delivery systems [9,10], and direct administration routes such as intracerebroventricular or intrathecal injection [11–13]. Considering this, understanding whether VAL-1221 can cross the BBB is critical to evaluating its potential as a CNS-targeting therapy.

Nowadays, MS—especially HRMS—coupled with electrospray ionization (ESI) and matrix-assisted laser desorption/ionization (MALDI) has become the predominant technique for bioanalysis. Advances in HRMS instrumentation, combined with LC, have significantly enhanced protein characterization and facilitated the ongoing development of biopharmaceutical products [14].

In recent years, several studies have highlighted the critical role of MS in analyzing, monitoring, and characterizing therapeutic proteins, especially biopharmaceuticals like mAbs and enzyme replacement therapies (ERTs) [15–18].

Taking advantage of the high potential of MS-based proteomics, this research aims primarily at the qualitative/quantitative characterization of the biotechnological drug VAL-1221 in plasma and CSF to verify the actual crossing of the BBB and, secondly, to expand the panorama of studies of biotechnological therapeutics by MS analytical techniques. To the best of our knowledge, this is the first study focused on the use of a micro liquid chromatography-high resolution mass spectrometry (microLC-HRMS/MS)-based proteomics method to assess the potential of VAL-1221 for Lafora disease.

## 2. Materials and methods

### 2.1 Chemicals and Reagents

VAL-1221 (10.0 mg/mL) was provided by Parasail LLC (Quincy, Massachusetts, USA). Sequencing Grade Modified Porcine Trypsin was obtained from Promega Corp (Madison, WI, USA). Hydrochloric acid, dithiothreitol (DTT), iodoacetamide (IAA), ammonium bicarbonate and HPLC-grade acetone were from Merck (Darmstadt, Germany). UHPLC-MS grade acetonitrile (ACN), UHPLC-MS grade methanol (MeOH) and water were provided by VWR chemicals (Radnor, PA, USA). LC-MS grade formic acid (FA) was purchased from Carlo Erba Reagents S.r.l. (Milan, Italy). Oasis^®^ HLB µElution Plate for solid phase extraction (SPE) was provided by Waters Corp. (Milford, MA, USA). Tris was obtained from BIO-RAD (Hercules, CA, USA). Guanidine Hydrochloride (Gua-HCl) was from PanReac AppliChem ITW Reagents (Glenview, IL, USA). Regenerated Cellulose (RC) Membrane 0.2 µm 4mm Syringe Filters were from Phenomenex (Torrance, CA, USA). SWATH Acquisition Performance Kit was provided by Sciex (Concord, Ontario, Canada). Radioimmunoprecipitation assay (RIPA) buffer and protease and phosphatase inhibitor cocktail were purchased from Thermo Fisher Scientific (Waltham, MA, USA).

### 2.2 Patients Enrollment

Ten patients with genetically confirmed LD, named LD01-LD10, at the mid-to-late stage of the disease, were enrolled at IRCCS Istituto delle Scienze Neurologiche di Bologna between May 2023 and June 2024. Five out of ten (LD02-LD06) were treated with VAL-1221 as part of a compassionate use program. VAL-1221 was administered according to the protocol described by Muccioli et al. [19]. Briefly, the liquid formulation (10.0 mg/mL) was diluted to 5.2 mg/mL in the infusion bags with physiological solution. The latter solution was administered to the patient intravenously at a dose of 20 mg/kg, starting with weekly infusions for three weeks, followed by infusions every other week. Patients underwent comprehensive assessments at baseline, at an intermediate time point, and at the end of the 1-year treatment to evaluate safety and exploratory efficacy endpoints, described in detail in a companion manuscript.

### 2.3 Bio-sample collection and storage

Plasma and CSF from LD-untreated patients (n=5) were collected and used as blank samples to prepare spiked samples, for qualitative and quantitative analytical aims.

Plasma samples from treated patients were collected to study the drug concentration profile at the following time points (min = minutes; h = hours): before infusion (t0), straight after the end of infusion (t1), after 30 min (t2), 1 h (t3), 4 h (t4), and 24 h (t5) post-infusion. CSF samples were also collected, and timing was tailored for each patient based on their plasma concentration profiles within 1 h and 24 h after the end of infusion. All biological samples were aliquoted and stored at −80 °C until analysis.

### 2.4 Stock and calibrating solutions

The VAL-1221 consists of a frozen liquid solution formulated at 10.0 mg/mL (68.05 µM) in 0.02 M Sodium citrate/citric acid, 0.15 M NaCl. Five Calibration solution levels were prepared in water from 68.05 µM stock solution: 6.8 (I), 68.0 (II), 340.0 (III), 680.0 (IV), 1700.0 (V) nM. Stock and calibration solutions were stored at −20 °C. The peptide reference solutions were obtained by following the digestion procedure described below. These solutions, containing intact or digested VAL-1221, were used to prepare spiked plasma/CSF and to determine validation parameters (see Section 2.5). VAL-1221 diluted solution in the infusion bags was also characterized.

### 2.5 Spiked plasma/CSF and samples from LD-treated patient solutions

Spiked CSF and plasma samples from LD-untreated patients were obtained by adding the VAL-1121 stock solution at concentration levels I to V.

The analysed biological samples from LD-treated patients were serum, plasma, peripheral blood mononuclear cells (PBMCs), CSF, and CSF pellets. PBMCs and CSF pellets (pre-processing protocols are detailed in the Supplementary materials) were resuspended in lysis buffer containing protease and phosphatase inhibitors in a 1:1 volume ratio. The Eppendorf tubes were placed on ice for 15 min, followed by two cycles of thermal shock (10 min at −20°C and 10 min at room temperature). Finally, samples were centrifuged at 14000 × g for 15 min, and the supernatant was processed using the protocol described below. Spiked and treated sample preparation for bottom-up proteomics followed the same procedure: 20 µL of biological sample was placed in a clean tube with 4 volumes of 20 mM NaCl/acetone (20/80 v/v), vortexed for 10 seconds, and left at room temperature (RT) for 2 min. After centrifugation at 14000 x g for 20 min, the supernatant was discarded, and the pellet was washed twice with neat acetone.

The dried pellet was resuspended in 50 µL of Gua-HCl 6M/Tris-HCl 50mM/DTT 5mM and incubated for 45 min at 56°C under gentle shaking. 7 µL of IAA 120 mM was added to the tubes to a final concentration of 15 mM and incubated again for 30 min at RT in the dark. Samples were diluted with 390 µL of Ammonium bicarbonate 50 mM, pH 7.8, and trypsin was added in a ratio of 1:50 enzyme: protein. Digestion took place overnight at 37°C under gentle shaking. Digestion was quenched by adding 50 µL of 10% formic acid to a final concentration of 1%.

After digestion, we used Oasis HLB µElution SPE plate wells for peptide sample clean-up. The cartridges were conditioned and equilibrated with 500 µL of MeOH, followed by 500 µL of 0.1% formic acid. After sample loading, each well was washed with 500 µL of 0.1% formic acid. Peptides were eluted with 100 µL of 80% acetonitrile/0.1% formic acid. Extracts were dried under vacuum at 37°C and resuspended with 100 µL of 0.1% formic acid. Each sample was filtered with a 0.2 µm RC syringe filter before injection.

The so-called post-spiked samples, used for recovery and matrix effect evaluation, were spiked with water solutions of digested VAL-1221 at a known concentration (see section 2.4) before microLC-HRMS analysis.

### 2.6 microLC-HRMS analysis

MS-based bottom-up proteomics involved the digestion of intact proteins (see Section 2.5) and the subsequent LC-MS analysis of the produced peptides. Analysis was performed on an Eksigent M5 MicroLC system (Sciex, Concord, Ontario, Canada) coupled to a TripleTOF 6600^+^ mass spectrometer with OptiFlow Turbo V Ion Source (Sciex, Concord, Ontario, Canada). 5 µL from each sample was loaded onto a Kinetex XB C18 50 × 0.5 mm I.D., 2.6 µm 100 Å. During the analysis, the column temperature was 35°C. Chromatographic separation occurred in 60 min with a constant flow rate of 20 µL/min. Gradient elution program was as follows: 0–40 min, 5-40% eluent B; 40-50 min, 40-60% eluent B; 50-51 min, 60-90% eluent B; 51-57 min, 90% eluent B; 57-58 min, 90-5% eluent B; 58-60 min, 5% eluent B. Equilibration time between chromatographic runs was 3 min. Mobile phase A consisted of 0.1% formic acid, and mobile phase B was acetonitrile/0.1% formic acid. IonSpray voltage (ISV) was 5000 V, and Curtain Gas supply pressure (CUR) was 30 PSI; nebulizer and heater gas pressures were set at 30 and 40 PSI, respectively. The ion spray probe temperature was 300 °C. Declustering potential was 80 V. Analyses were carried out using rolling collision energy mode.

Samples were analysed in Information Dependent Acquisition mode (IDA). PepCal Mix (Sciex, Concord, Ontario, Canada) was used to ensure steady MS and MS/MS calibrations throughout the analysis.

Quantitative analysis was performed on the extract ion chromatogram (EIC) peak area generated by off-line extrapolation of the signals at m/z 553.85 and 461.71, corresponding to multi-protonated molecular ions of the Peptide Containing-Tag (PC-Tag) sequence, [PC-tag+5H]^5+^ and [PC-tag+6H]^6+^, respectively.

### 2.7 Method validation

The validation was performed using EMA guidelines for bioanalytical methods, and the following parameters were tested [20,21]. All the calculations were performed by considering the PC-Tag peptide peak area obtained from extract ion chromatograms.

#### Linearity

water and spiked plasma/CSF calibrators were prepared at the concentration levels I-V (see Section 2.4). The peak areas of the PC-tag response in EIC ([tag+5H]^5+^ at 553.85 m/z) were plotted against the nominal concentrations. The calibration curves were set up using a linear least-squares regression method. Linearity was assessed by determining the coefficient of determination (R^2^). The acceptance requirement was R^2^ ≥ 0.996. The back-calculated concentrations of the calibration standards must be within ±15% of the nominal value.

#### Lower limit of detection (LLOD)

LLOD was determined experimentally in triplicate by comparing measured signals from spiked samples at decreasing concentrations (serial dilutions) with those of blank plasma/CSF at the PC-tag corresponding retention time. LLOD was calculated as 3 times the baseline noise, i.e., signal-to-noise ratio (S/N) equals 3.

#### Lower limit of quantification (LLOQ)

LLOQ was experimentally defined as the lowest quantifiable concentration of analyte in spiked samples, with acceptable accuracy and precision (≤ ±20%) and a S/N of 10. LLOQ was estimated, as for LLOD, by analyzing plasma/CSF sample spiked solutions at serial dilution concentrations, based on the criteria of accuracy and precision.

#### Selectivity

was evaluated through injections of blank plasma and CSF matrices. The method was considered selective and accepted if the peak area of the PC-tag extract ion chromatograms was less than 20% of the LLOQ.

#### Precision and accuracy

intra- and inter-day precision and accuracy were assessed by determining the coefficient of variation (CV%) and relative error (RE%), respectively.

Intraday precision and accuracy were evaluated by analyzing six spiked plasma and CSF samples at concentrations II, III, and IV. Interday precision and accuracy were assessed by analyzing the same spiked samples from three different runs on three different days. The acceptance criteria should not exceed 15% CV for the spiked samples, except for the LLOQ, which should not exceed 20%.

#### Recovery

the recovery (R%) was determined as the percent ratio between the mean (n = 3) of the peak areas of the analyte in spiked plasma/CSF samples (before digestion, named pre-spiked samples) and those obtained by spiking the post-digestion solutions with digested VAL-1221 (post-spiked samples) at the same VAL-1221 original concentration. Pre- and post-spiked plasma/CSF samples were prepared at II, III, and IV concentrations.

#### Matrix effect

was investigated in triplicate, using plasma and CSF from untreated patients spiked with digested VAL-1221 (post-spiked samples) at concentration III. The matrix factor (MF%) was calculated as a ratio between the PC-tag area in spiked samples and standard analyte solutions. The MF% should be between 80 and 120.

#### Carry-over

was performed by injecting a blank sample after the highest calibrator. Carry-over was considered negligible if it was ≤ 20% than LLOQ.

#### Stability tests

to assess the stability of the digested samples (spiked plasma/CSF), we performed the analysis after storing the solutions at II, III, and IV concentration levels in the autosampler (T = 4 °C) for 48 h and at −20 °C for 15 days. Stability expressed as percentage change (%C) was calculated by comparing the concentration before and after storage as described in equation (1). Samples were considered stable if their mean concentrations were within ±15% of the nominal concentration.

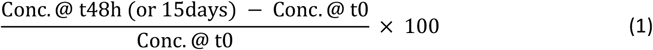

### 2.8 VAL identification and concentration profile of LD-treated patients

Plasma and CSF samples from treated patients were analysed to confirm the VAL-1221 presence by its identification. VAL-1221 in plasma was quantified at the time points t0, t1, t2, t3, t4, t5, differently in CSF at t1, t3, t5. To assess drug distribution in different parts of the biospecimens, in addition to plasma and CSF, we analysed serum, PBMCs, and CSF pellets from treated and LD-untreated patients.

All biological samples were stored at −80 °C until analysis. Biological samples were processed and analyzed as described above (see sections 2.5 and 2.6).

### 2.9 Data analysis for protein identification

We processed the obtained data with ProteinPilot software (Sciex, Concord, Ontario, Canada) with trypsin set as the digestion enzyme and carbamidomethylation (CAM) as a constant modification for cysteine. Matching was carried out against the UniProt Human Proteome (downloaded in August 2023) database using the Paragon algorithm, modified by the addition of the drug sequence. Biological modification mode was enabled to account for variable modifications of peptides. False Rate Discovery analysis was performed, and proteins with 95% confidence (Unused score ≥ 1.3) were identified.

## 3. Results

We analyzed biological samples with a bottom-up proteomic approach, both in untargeted, for identification, and targeted modes. Targeted quantitation of VAL-1221 was carried out by monitoring a unique peptide containing the TAG sequence, which was introduced, as a linker, during the biotechnological drug VAL-1221 preparation process to fuse the cell-penetrating Fab antibody fragment with recombinant human acid alpha-glucosidase to facilitate intracellular enzyme delivery. The predicted 3D structure, and the scheme of the VAL-1221 are reported in Figure 1.

**Fig. 1.**
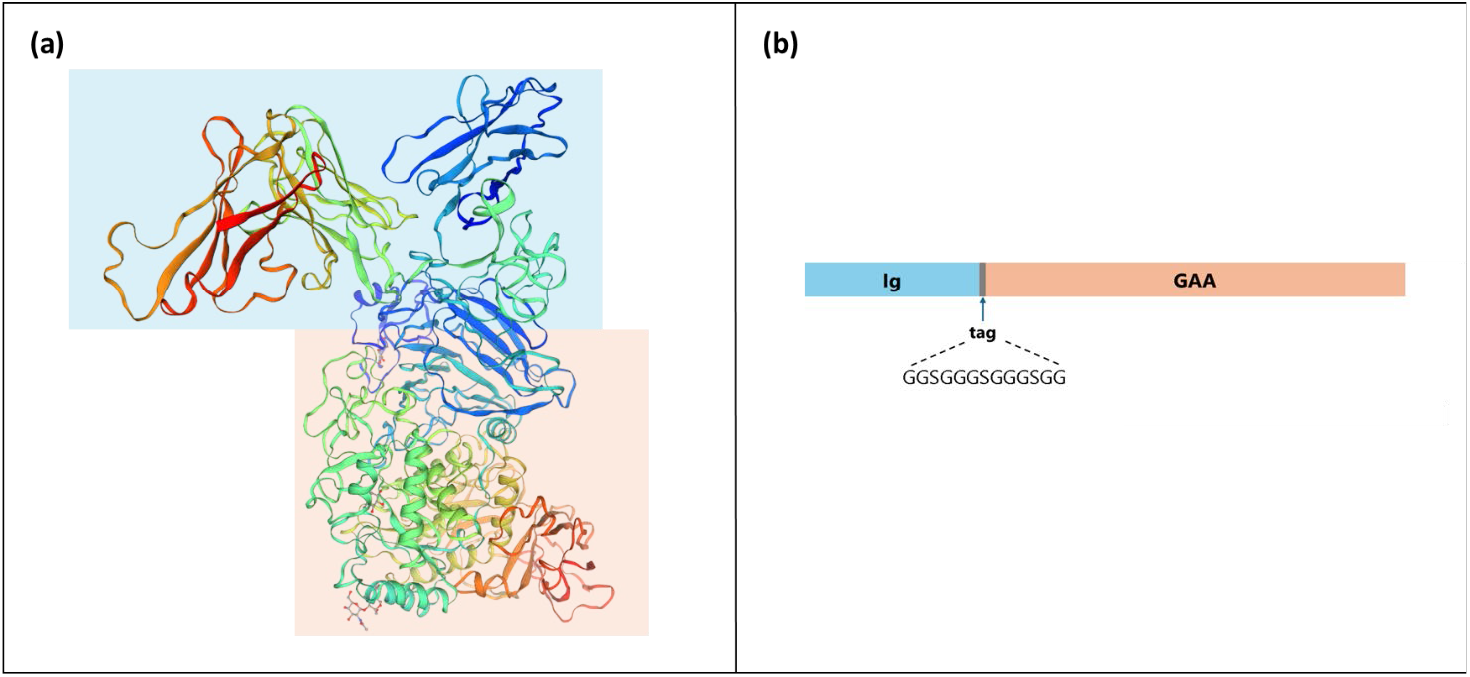
Structural representation of the biotechnological drug VAL-1221. (a) The predicted three-dimensional (3D) structure was generated by homology modeling using a template-based approach via the SWISS-MODEL platform (https://swissmodel.expasy.org/docs/references) [22]. (b) VAL-1221 2D scheme. The Fab fragment of the hu3E10 antibody is shown in blue, including both the heavy and light chains. The light chain is disulfide-bonded to the Fd portion of the heavy chain, which is genetically fused to residues 67-952 of the human acid α-glucosidase (GAA) enzyme, shown in orange.

To assess the feasibility of the analytical method, the approach of this study initially involved the characterization of VAL-1221 in the aqueous solution of the pharmaceutical formulation (10 mg/mL), followed by its identification in the infusion bag (5.2 mg/mL) and in spiked biological matrices (Figure 2). The next step was the development and validation of a quantitative method based on the PC-tag signal. Finally, the study assessed the presence of the biotechnological drug in plasma at different time points post-administration and in the CSF, to study the possible crossing of the BBB.

**Fig. 2.**
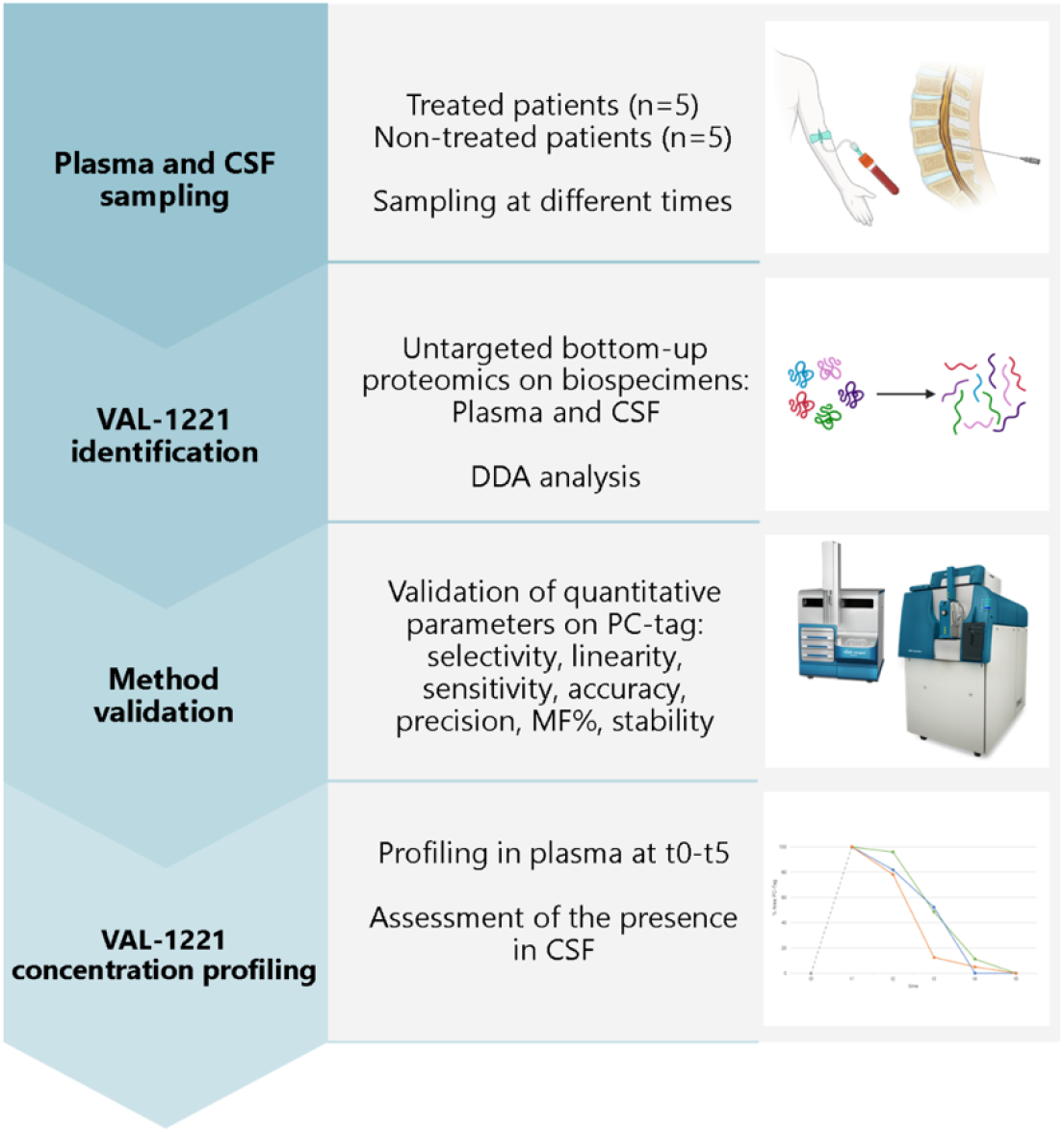
Methodological workflow of the study. Plasma and cerebrospinal fluid (CSF) samples were collected from treated (n=5) and untreated (n=5) patients at multiple time points. VAL-1221 was identified on spiked plasma and CSF samples from untreated patients using untargeted bottom-up proteomics, followed by data-dependent acquisition (DDA) analysis. Method validation was performed based on the PC-tag, evaluating key parameters including selectivity, linearity, sensitivity, accuracy, precision, matrix factor (MF%), and stability. Finally, VAL-1221 concentration profiling on treated patients was conducted in plasma at time points t0–t5, and its presence was assessed in CSF.

### 3.1 Untargeted Proteomics: VAL-1221 identification in plasma and CSF

A bottom-up untargeted MS-based proteomics approach was employed for drug identification, focusing on peptide fragmentation profiling. After the identification of VAL-1221 in the pharmaceutical formulation and infusion bag, these data were used to confirm the identification in biospecimens.

#### 3.1.1 Spiked samples

Biological samples were processed using standard procedures for protein extraction, enzymatic digestion, and peptide cleanup as described in section 2.5.

After microLC-HRMS/MS analysis, the data were then processed using ProteinPilot software. It applies algorithms to match the observed peptide masses with the reference protein UniProt Human Proteome database, modified by the addition of the drug sequence. This approach allowed for the accurate identification of hundreds to thousands of proteins in plasma and CSF. Among these proteins, we identified VAL-1221 with an untargeted approach within the entire human proteome database, ensuring a non-hypothesis-driven identification. The sequence of identified peptides, with coverage percent (%Cov), unused values, and sequence are reported in Table S1.

Table 1 reports information on VAL-1221 identification (number of identified peptides, %Cov, and unused values) and microLC-HRMS analytical data (retention time, m/z values, and peptide charges) obtained for PC-tag.

**Table 1.**
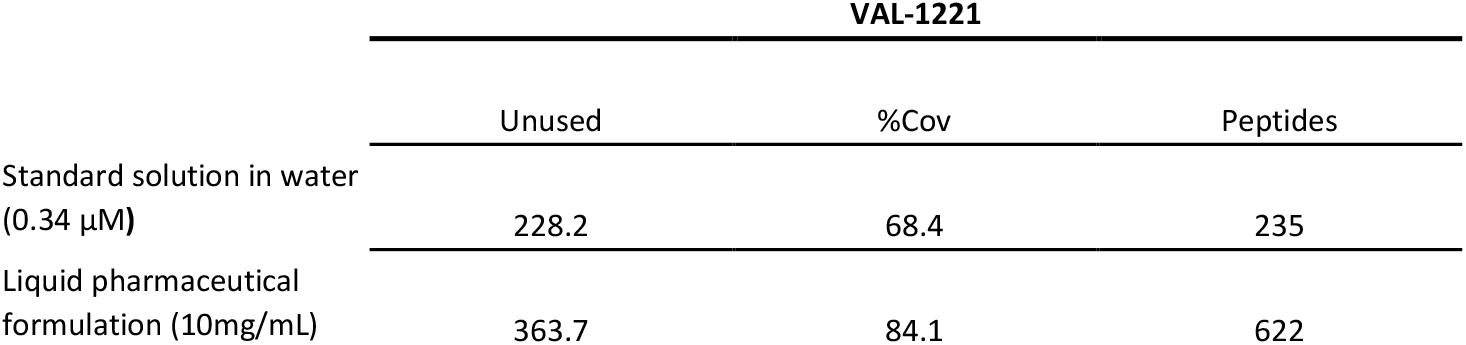
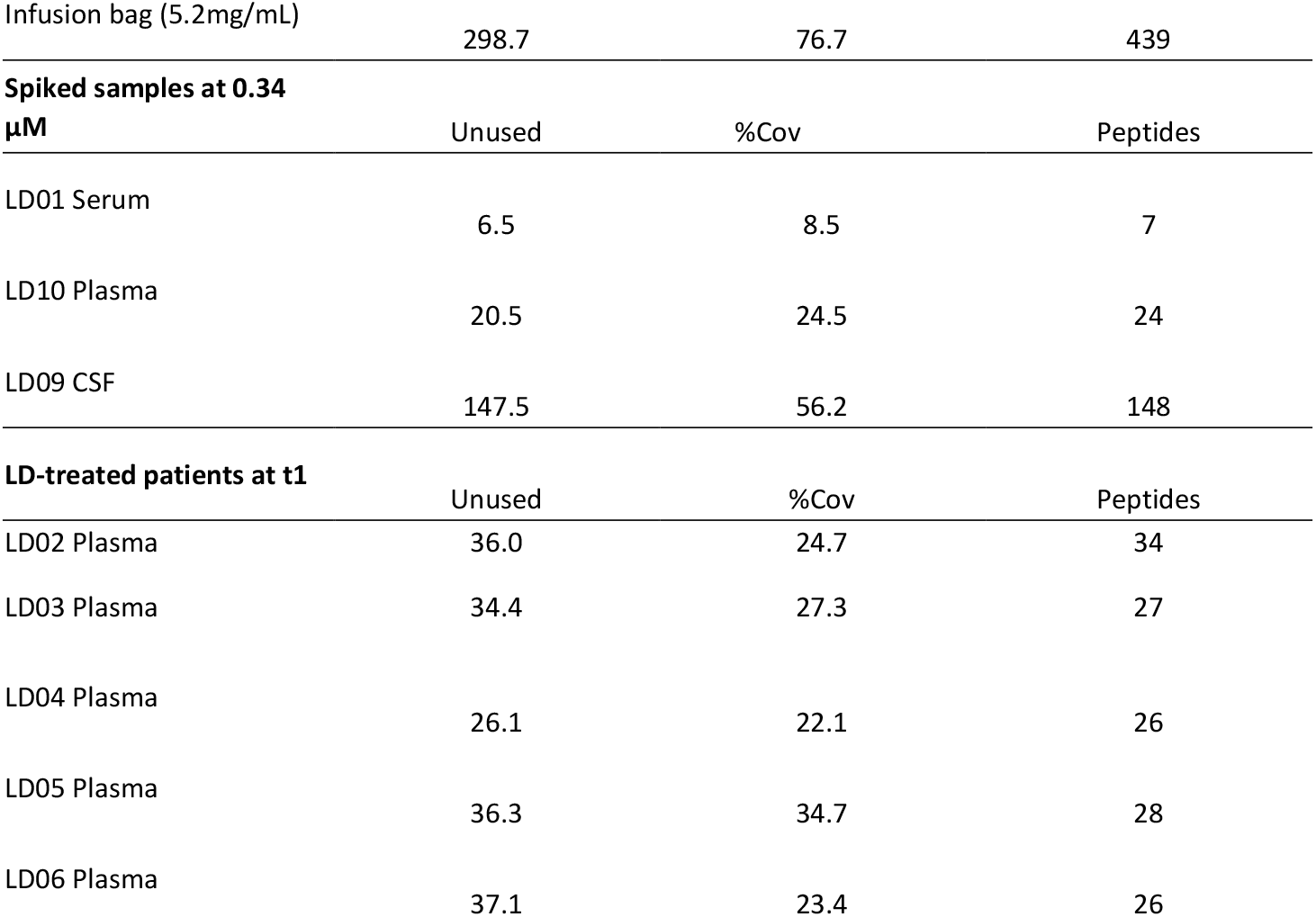
Identification parameters of VAL-1221 in standard solution, pharmaceutical formulation, infusion bag, spiked samples (serum, plasma, CSF), plasma of LD-treated patients: number of unused peptides, percentage of protein coverage (%Cov), number of identified peptides. Confidence level 95%.

VAL-1221 was analyzed in different spiked matrices to compare the number and positions of tryptic cleavages, peptide coverage, and unused score, to validate the identification. Table 2 shows the identification parameters and the number of the identified peptides in standard aqueous solution (0.34 µM), liquid pharmaceutical formulation (vials), infusion bags, and 0.34 µM spiked serum/plasma/CSF.

#### 3.1.2 LD patient samples

We analyzed a comprehensive panel of biological samples collected from LD patients at multiple time points following drug infusion. Serum, plasma, and CSF samples were obtained from treated and untreated individuals at different time points as reported in Table S2.

The drug was detected exclusively in plasma samples from treated patients, and only up to time point t4. No detectable levels were observed in serum or CSF, nor in any samples from untreated individuals.

VAL-1221 identification parameters for plasma and CSF samples of treated patients are reported in Table 1.

### 3.2 Targeted Proteomics: VAL-1221 quantitation

The PC-tag sequence was targeted for a very selective quantitative monitoring purpose.

The PC-Tag multicharged ions, [tag+5H]^5+^, at 553.85 m/z and a retention time of 4.7 min, were monitored as quantifier ions (see Section 2.6). The peak areas of the extracted ion chromatograms (Figure 3) were used for VAL-1221 absolute quantitation, correlating it with the calibration curve obtained in the same matrix. The [tag+6H]^6+^ signal at 461.71 was used as qualifier ion.

**Fig. 3.**
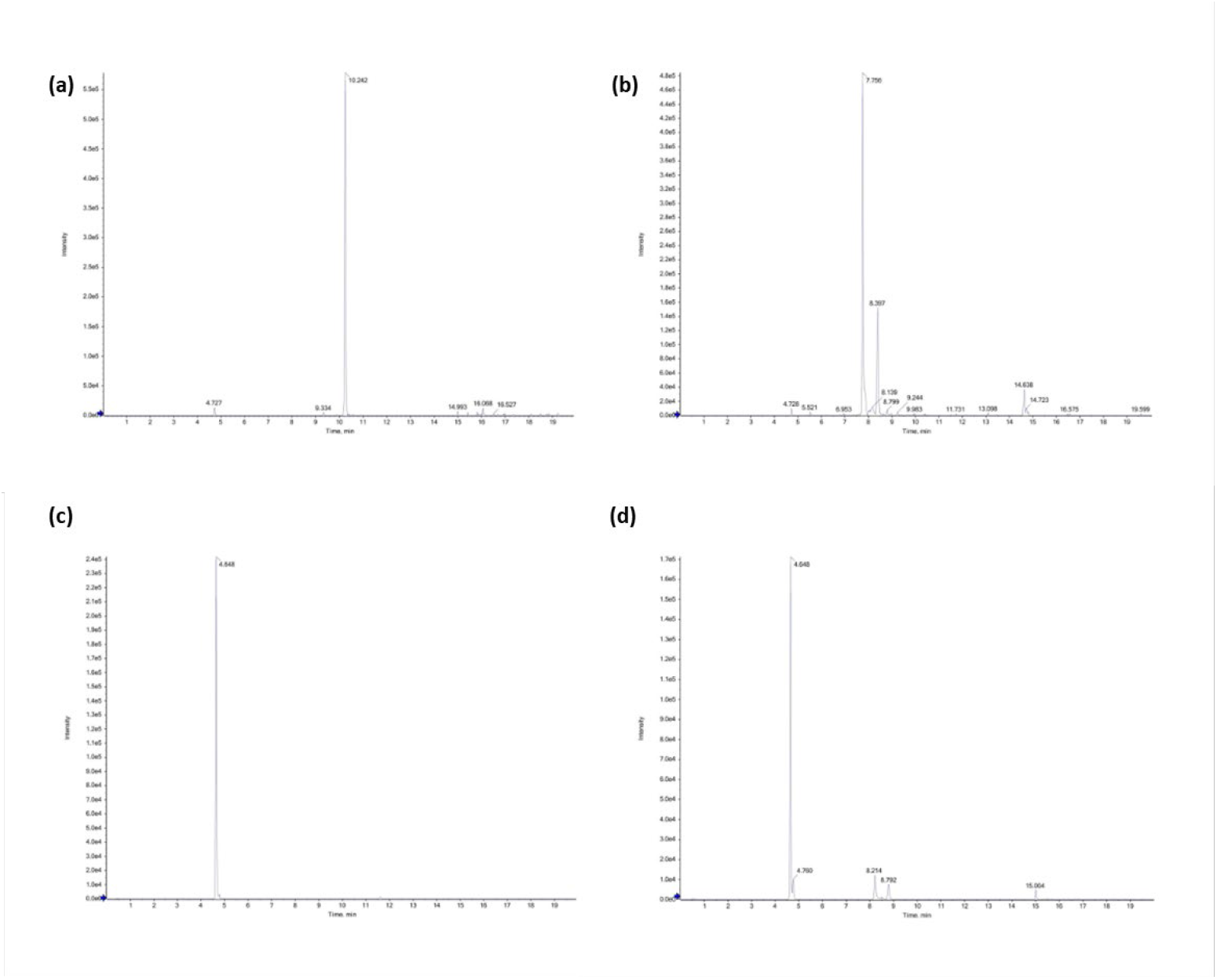
XIC chromatogram of spiked samples at 340 nM: (a) quantifier ion (m/z 553.850 ± 0.005) and (b) qualifier ion (m/z 461.710 ± 0.005) in plasma sample, (c) quantifier ion (m/z 553.850 ± 0.005) and (d) qualifier ion (m/z 461.710 ± 0.005) in CSF sample.

### 3.3 Method validation

The optimized analytical conditions were validated according to the EMA guidelines for bioanalytical methods, and the parameters described in Section 2.7 were evaluated. The method was proved to be linear and selective for the whole concentration range studied (see Table 3 and blank chromatograms in Figure S1 of the supplementary material, respectively). The results for intraday (n = 6) and interday (over 3 days) precision and accuracy are presented in Table 3, demonstrating satisfactory performance. Intraday precision and accuracy were within ±15%, with acceptable %RSD (≤15) and %RE (≤15). Interday precision and accuracy were within ±15%, with RSD% ranging from 10.3 to 14.7 in the different matrices. Table 3 also includes LLOQ concentrations.

**Table 3.**
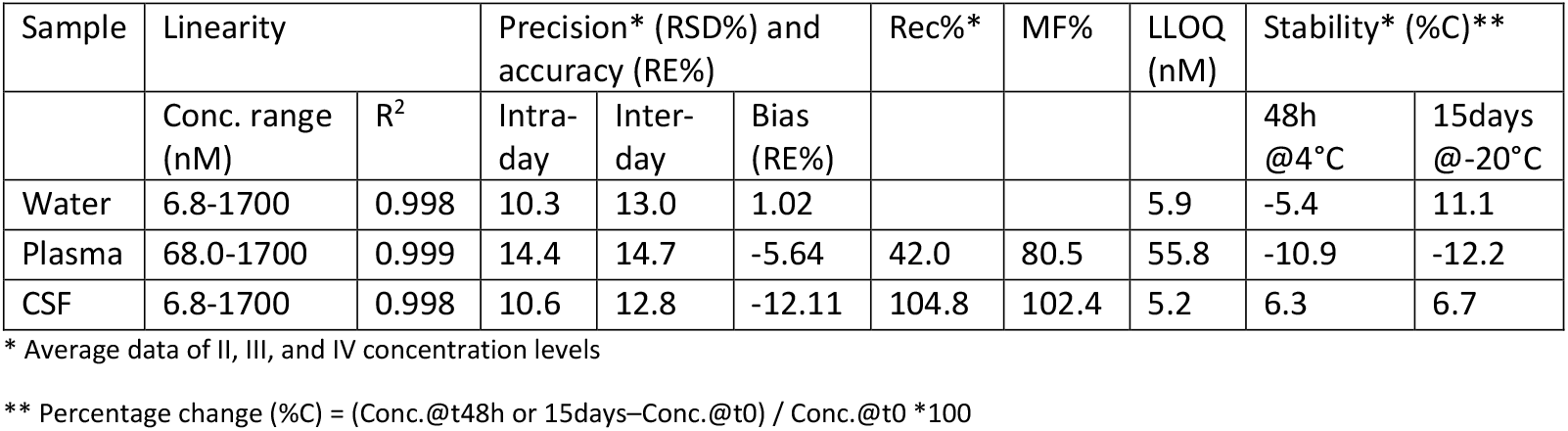
Method validation parameters.

The matrix factor in plasma/CSF samples was evaluated on 3 different blanks from untreated LD patients. After protein tryptic digestion, samples were spiked with digested VAL-1221 at 0.68 µM (post-spiked). The signal in the post-spiked sample was compared with those in the aqueous solutions at the same concentration. The average matrix factor was found to be between 80 and 102% in the two matrices (all data are represented in Table 3). The values showed that the matrix effect is negligible (within ±20%) in CSF; on the other hand, the matrix effect in plasma is higher, although it remains below 20%.

The recovery was calculated by comparing pre- and post-spiked plasma/CSF samples. The results, as Recovery% (Rec%), shown in Table 3, were satisfactory for CSF; on the contrary, in plasma, the recovery was low, in the order of 40%. In an attempt to understand this low recovery, we excluded the possible internalization of VAL-1221 by PBMC, which would remove the drug from the plasma after centrifugation. The VAL-1221 content determination in PBMCs, by PC-tag quantification, proved its absence and excluded compartmentalization (data not shown).

LLOD and LLOQ values (in Table 3) reflected matrix effect and recovery, being much lower in CSF. LLOD ranged from 2 nM in water and CSF to 30 nM in plasma. Carry-over was found to be ≤ 20% than LOQ. All criteria based on EMA guidelines for method development were satisfied in CSF.

VAL-1221 standard and spiked solutions were evaluated as stable after 2 days in an autosampler at 4°C and after storage at −20°C for 15 days, with a concentration change within ±15%. (Table 3)

### 3.4 Evaluation of the presence of VAL-1221 in plasma and CSF after intravenous administration

Due to the linear relationship between signal intensity of the PC-tag and the abundance of the target protein, we were able to quantify VAL-1221 in plasma samples from 3 treated subjects at different time points. The results are reported in Figure 4.

**Fig. 4.**
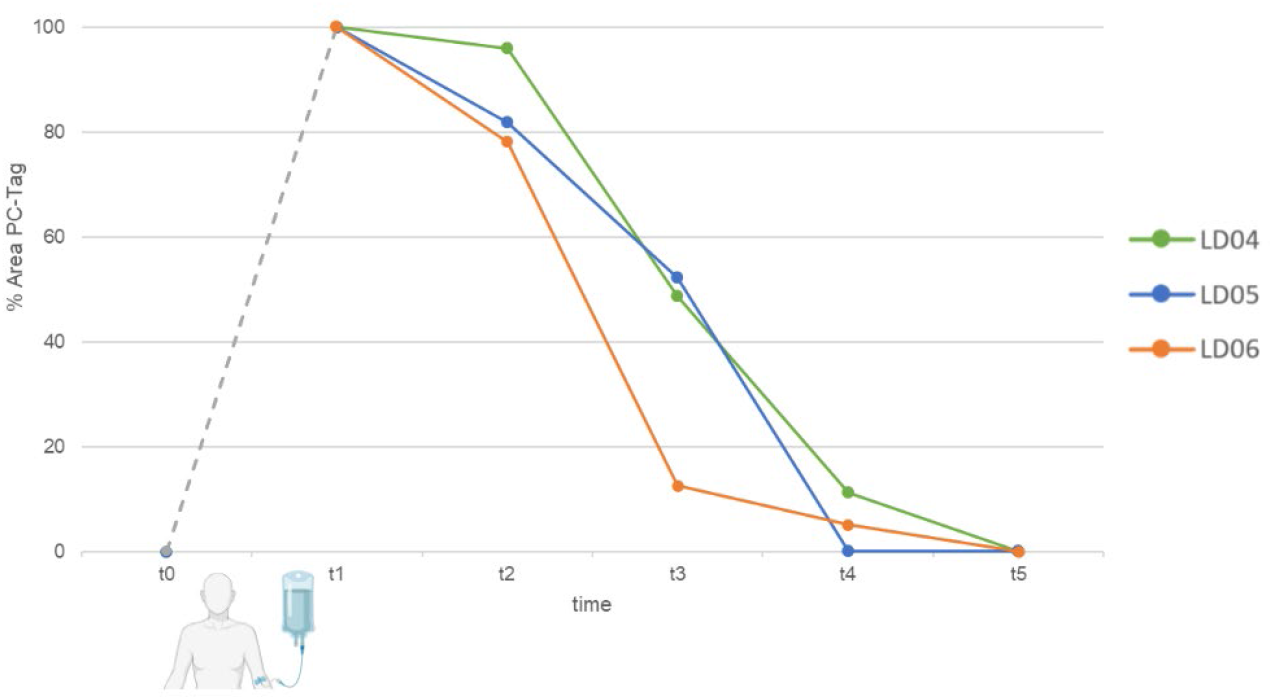
PC-tag concentration (%) profile of VAL-1221 in plasma from three LD-treated patients. Time points include: t0 (pre-dose), t1 (end of infusion), t2 (30 min post-infusion), t3 (1 h post-infusion), t4 (4 h post-infusion), and t5 (24 h post-infusion). The percentage values are normalized to t1 (100%), which corresponds to the end of the intravenous infusion. The infusion duration ranged from 2.5 to 3 h.

Extract ion MS analysis produced reproducible results showing a significant decreasing trend in VAL-1221 concentration from the end of administration (duration 2.5-3 h), defined as time 1 (t1), after 30 min (t2), 1 h (t3), 4 h (t4), and 24 h (t5). As shown in Figure 3, no detectable drug traces were found 24 h after the end of administration. The amount of VAL-1221 was plotted as a percentage ratio between the PC-tag area at each time point after administration (t2-5) and the PC-tag area at t1.

The concentration of VAL-1221 at t1, estimated using the multiple standard additions method, ranged from 0.68 to 1.02 µM. These values are lower than the theoretical concentration of 1.77 µM, calculated assuming an average blood volume of 5 L and no drug distribution.

Based on this assumption, CSF was analysed right after 1h and 24h after the end of the infusion. In CSF samples, despite the presence of a few peptides attributed to the endogenous antibody, the PC-tag signal was not detected, nor were the enzyme-related peptides. VAL-1221 was not detected in serum, PBMCs, and CSF pellets.

## 4 Discussion

Thanks to the optimization and validation of the method based on high-resolution mass spectrometry, we were able to identify, characterize, and quantify VAL-1221 and to profile its concentration in plasma after administration in patients with LD. Notably, VAL-1221 is the first potential disease-modifying therapy investigated in LD, a devastating neurodegenerative disorder that currently has no cure.

The determination was selective and sensitive thanks to the use of the synthetic tag as a chemical labeling. In general, a peptide tag is a short, synthetic peptide sequence that is added to a protein or peptide of interest to facilitate its identification and quantification in mass spectrometry-based proteomics. The tag acts as a unique marker that can be easily detected and distinguished from the endogenous peptides in the sample.

Method validation demonstrated high precision, accuracy, good sensitivity for CSF, and stability under storage conditions (RSD%≤15, RE%≤15, Table 3).

The results showed the presence of VAL-1221 in the plasma of LD-treated patients; on the contrary, it was not identified in serum, PBMCs, CSF, and CSF-cells samples from treated subjects. Analysis of PMBCs and CSF pellets was conducted to investigate the possible distribution of the drug into other compartments, due to cellular uptake or precipitation, in an attempt to explain the low recovery of VAL-1221 in plasma and its absence in CSF. Regarding the serum, it can be inferred that VAL-1221 precipitates during clotting with the cellular components of the blood. Considering our results, we can assert that the inability to detect it in the PBMCs, cerebrospinal fluid, and CSF-cells of treated subjects may be attributed to either its low concentration (below the method LLOD) or its actual absence. We can also reasonably hypothesize that VAL-1221 is distributed in districts other than blood, and the analyzed bio-samples, concomitantly with the administration. Unfortunately, to date, validated studies monitoring biotech drugs in the CSF are very few [23], making it impossible to compare the sensitivity levels of our method.

Together with a low recovery, the matrix effect in plasma was higher than in CSF. This may be due to ionic suppression in ESI by the more complex matrix than CSF, as well as the higher concentration of interfering endogenous proteins. Due to the high matrix effect in plasma and low recovery, the actual concentration values in this matrix should be corrected by the ME% and Rec%.

After applying the correction factor ME%-Rec%, the blood concentration profile showed a decrease in VAL-1221 levels over time and a concentration at t1 lower than the intended dose, suggesting that the drug is distributed out of the bloodstream soon after administration and before its termination.

Although the drug was present in systemic circulation, these findings indicate limited penetration through the blood-CSF barrier, despite the low LLOD of the analytical method and the effect of pre-concentration of the CSF sample by SPE. Our results could pose a challenge to its therapeutic efficacy for LD, particularly regarding CNS manifestations. As preclinical data have demonstrated clearance of Lafora bodies from the brain following intracerebroventricular delivery of VAL-1221, alternative administration routes may represent a more promising strategy to overcome the blood–brain barrier. Such approaches, while technically and logistically challenging, could be crucial for future efforts to achieve disease modification in LD.

Beyond their implications for LD, this analytical approach may also have translational value for other glycogen storage disorders. Since VAL-1221 was originally developed for Pompe disease, understanding its distribution, recovery, and limitations in human samples can inform future clinical development and guide the design of optimized delivery strategies in that population as well.

## 5 Conclusion

In the limited literature on validated studies monitoring biotech drugs in the CSF, this study represents the first investigation of the potential use of the biotechnological drug VAL-1221 for the treatment of Lafora disease. In a compassionate study in which VAL-1221 was administered intravenously to LD patients, we assessed the possible central distribution of the drug by analyzing CSF using MS-based proteomics. To this end, we optimized and validated the analytical method on spiked bio-samples to identify the drug in plasma and CSF in an untargeted approach and quantify it accurately and precisely by a targeted approach.

From an analytical point of view, the study demonstrates the growing importance of MS-based techniques in the characterization of biopharmaceuticals. MS not only helps analyze protein structure and modifications but also provides crucial insights for optimizing the production, stability, and effectiveness of therapeutic proteins.

The results demonstrated that the developed mass spectrometry method was able to monitor VAL-1221 in plasma after administration, and that it was not detectable in CSF at different time intervals post-administration. These findings raise the possibility that VAL-1221 does not effectively cross the blood–brain barrier or reach therapeutic concentrations in the CNS under the current treatment conditions. Notably, no clinical improvement was observed in LD-treated patients over 12 months of compassionate-use administration, as reported in a preprint study by our group. This does not preclude either the effectiveness of the drug itself in cleaning the LBs, nor the possibility of studying different administration routes or more massive doses that are still tolerable for patients.

## Data Availability

All data produced in the present study are available upon reasonable request to the authors

## Supplementary Information

The online version contains supplementary material.

## Acknowledgements

The study drug and amino acid sequence of VAL-1221 were supplied by Parasail LLC (CEO Dustin Armstrong).

## Declarations

### Funding

This study was funded by the European Union - Next Generation EU - NRRP M6C2 – Investment 2.1 Enhancement and strengthening of biomedical research in the NHS. Project ‘Drug discovEry and repurposing to Find a trEAtmenT for Lafora Disease (DEFEAT-LD)’ - PNRR-MR1-2022-12376430 and supported by the “Ricerca Corrente”, funding from the Italian Ministry of Health.

### Conflicts of interest

no conflict of interest

### Ethics approval

The local ethics committee “Comitato Etico Area Vasta Emilia Centro” approved the compassionate use program of intravenous VAL-1221 for the five patients with LD (approvals no. 19, 20, 21, 22, 23-2023-COMPASS-AUSLBO) and the proteomics study (protocol number 178-2024-SPER-AUSLBO) in both treated and untreated subjects.

### Consent to participate

All patients and/or their legal representatives gave informed written consent for participation.

### Consent for publication

not applicable

### Availability of data and material

All data produced in the present study are available upon reasonable request to the authors.

### Code availability

not applicable.

### Author contributions

Erika Esposito: Formal MS analysis, Data analysis, Visualization, Writing original draft, review & editing. Alice Caravelli: Formal MS analysis, Writing original draft, review & editing. Lorenzo Muccioli: Conceptualization, Patient enrollment, Writing – review & editing. Chiara Cancellerini: Writing – review & editing. Maria Tappatà: Patient enrollment. Eleonora Pizzi: Samples preprocessing, Writing. Raffaela Minardi: Samples preprocessing, Writing. DEFEAT-LD Study Group: funding acquisition, review & editing Valerio Carelli: Review & editing. Luca Vignatelli: Review & editing. Roberto Michelucci: Conceptualization, Writing - review & editing. Francesca Bisulli: Conceptualization, Writing – review & editing, Funding acquisition. Jessica Fiori: Conceptualization, Methodology, Supervision, Writing original draft, review & editing, Project administration, Funding acquisition.

### Open Access

yes

## APPENDIX

### DEFEAT-LD STUDY GROUP: LIST OF COLLABORATORS

Cosimo Altomare^1^; Massimo Carella^2^; Cinzia Costa^3,4^; Giuseppe Damante^5,6^; Lidia Di Vito^7^; Giuseppe d’Orsi^2^; Nicola Gambacorta^2^; Paola Imbrici^1^; Valentina Imperatore^3,8^; Antonella Liantonio^1^; Laura Licchetta^7^; Raffaele Lodi^7^; Paola Mantuano^1^; Serena Mazzone^7^; Orazio Palumbo^2^; Elena Pasini^7^; Paolo Prontera^8^

^1^ Dipartimento di Farmacia-Scienze del Farmaco, Università degli studi di Bari Aldo Moro, Bari, Italy

^2^ Fondazione IRCCS-Casa Sollievo della Sofferenza, San Giovanni Rotondo, Italy

^3^ Department of Medicine and Surgery, University of Perugia, Perugia, Italy

^4^ Section of Neurology, S. Maria della Misericordia Hospital, Perugia, Italy

^5^ Department of Medicine (DAME), University of Udine, Udine, Italy

^6^ Institute of Medical Genetics, Udine University Hospital, Udine, Italy

^7^ IRCCS Istituto delle Scienze Neurologiche di Bologna, Full Member of the ERN EpiCARE, Bologna, Italy

^8^ Medical Genetics Unit, S. Maria della Misericordia Hospital, Perugia, Italy.

## Notes

### Competing Interest Statement

The authors have declared no competing interest.

### Author Declarations

The local ethics committee Comitato Etico Area Vasta Emilia Centro (CE-AVEC) approved the compassionate use program of intravenous VAL-1221 for the five patients with LD (approvals no. 19, 20, 21, 22, 23-2023-COMPASS-AUSLBO) and the proteomics study (protocol number 178-2024-SPER-AUSLBO) in both treated and untreated subjects.

