## supplementary for "Assessment of the potential use of VAL-1221 for Lafora disease: MS-based proteomics for the characterization and quantitation of the biotechnological drug in plasma and cerebrospinal fluid"

[1] IRCCS, Istituto delle Scienze Neurologiche di Bologna, Laboratorio di Proteomica Metabolomica e Chimica Bioanalitica, Bologna, Italy
[2] IRCCS Istituto delle Scienze Neurologiche di Bologna, Full Member of the European Reference Network for Rare and Complex Epilepsies (EpiCARE), Bologna, Italy
[3] Department of Biomedical and Neuromotor Sciences, University of Bologna, Bologna, Italy
[4] Department of Chemistry "G. Ciamician", University of Bologna, Italy
§DEFEAT-LD study group: a list of collaborators is provided at the end of the manuscript as APPENDIX

***Corresponding author**:
Lorenzo Muccioli, MD, PhD
IRCCS Istituto delle Scienze Neurologiche di Bologna
Via Altura 1/8, 40139 Bologna - Italy

ORCID: 0000-0001-6827-0099

**Pre-processing protocols.** Peripheral blood was collected from all participants after overnight fasting, using EDTA tubes for PBMC and plasma isolation, and serum separator tubes for serum collection.

For PBMC isolation, blood was diluted 1:1 with PBS (1×), layered onto Lympholyte separation medium (Lympholyte-H, Cedarlane), and centrifuged at 800 x g for 20 minutes at room temperature. The PBMC layer was carefully aspirated, washed with PBS, and centrifuged at 3200 x g for 10 minutes at 4 °C. The pellet was resuspended in 2 mL PBS(1x), then erythrocyte lysis was performed by adding 8 mL of $NH_4Cl$ solution (1x) and incubating for 10 minutes at room temperature. After an additional centrifugation at 3200 x g for 10 minutes at 4 °C, the pellet was resuspended in 2 mL PBS (1×), aliquoted into vials, and centrifuged once more at 4300 x g for 10 minutes at 4 °C. Cell pellets were either stored as dried PBMCs.

Serum was obtained by allowing the blood to clot for 1 hour, followed by centrifugation at 3200 x g for 10 minutes at 4 °C.

Plasma was processed within 1 hour of collection by sequential centrifugations at 3200 x g and 4300 x g, both for 10 minutes at 4 °C, and aliquoted prior to storage.

CSF samples underwent an initial centrifugation at 3200 x g for 10 minutes at 4 °C to separate the cell-free fraction from the cellular pellet. The cell-free fraction was clarified by centrifugation at 4300 x g for 10 minutes at 4 °C, while the pellet was resuspended in 50 µL of residual CSF, centrifuged at 4300 x g for 10 minutes at 4 °C, and then the supernatant was discarded.

All samples were stored at −80 °C until analysis.

**Table S1** Identification parameters of VAL-1221 Heavy Chain-GAA fusion in water and spiked plasma/CSF: number of unused peptides, percentage of protein coverage (%Cov), and number of identified peptides at a confidence level of 95%.

| **$H_2O$ 0.34 µM** | | |
|---|---|---|
| *VAL-1221 Heavy Chain-GAA fusion* | | |
| Unused | %Cov | Peptides |
| 228,2 | 68,4 | 235 |

| **Plasma 0.34 µM** | | |
|---|---|---|
| *VAL-1221 Heavy Chain-GAA fusion* | | |
| Unused | %Cov | Peptides |
| 20,5 | 24,5 | 24 |

| **CSF 0.34 µM** | | |
|---|---|---|
| *VAL-1221 Heavy Chain-GAA fusion* | | |
| Unused | %Cov | Peptides |
| 147,5 | 56,2 | 148 |

**Table S2** Overview of sample collection from LD-treated (t) and LD-untreated (ut) patients. Serum, plasma, and cerebrospinal fluid (CSF) samples were collected from ten individuals, identified as LD01 to LD10, at the following time points: before infusion (t0), immediately after the end of infusion (t1), 30 minutes post-infusion (t2), 1 hour (t3), 4 hours (t4), and 24 hours post-infusion (t5).

| | Serum | | | | | | Plasma | | | | | | CSF | | | | | |
|---|---|---|---|---|---|---|---|---|---|---|---|---|---|---|---|---|---|---|
| Time points | 0 | 1 | 2 | 3 | 4 | 5 | 0 | 1 | 2 | 3 | 4 | 5 | 0 | 1 | 2 | 3 | 4 | 5 |
| LD01 – ut | | | | | | | | | | | | | | | | | | |
| LD02 - t | | | | | | | | | | | | | | | | | | |
| LD03 - t | | | | | | | | | | | | | | | | | | |
| LD04 - t | | | | | | | | | | | | | | | | | | |
| LD05 - t | | | | | | | | | | | | | | | | | | |
| LD06 - t | | | | | | | | | | | | | | | | | | |
| LD07 - ut | | | | | | | | | | | | | | | | | | |
| LD08 - ut | | | | | | | | | | | | | | | | | | |
| LD09 - ut | | | | | | | | | | | | | | | | | | |
| LD10 - ut | | | | | | | | | | | | | | | | | | |

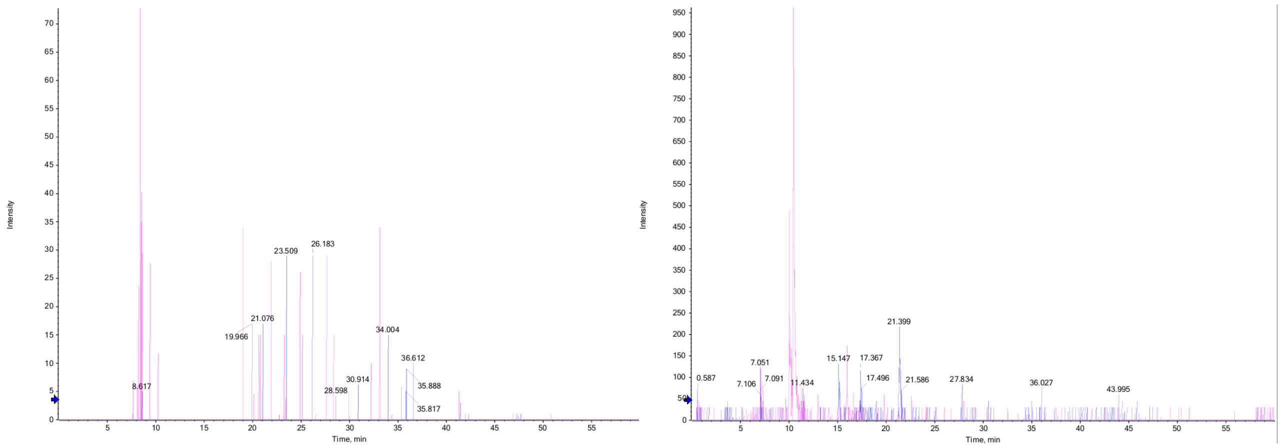


**Fig. S1** XICs from plasma (left) and CSF (right) samples of patient LD09 at 50 µg/mL: blue traces correspond to m/z 553.850 ± 0.005 Da, pink traces to m/z 461.710 ± 0.005 Da.
